# SARS-CoV-2 transmission in intercollegiate athletics not fully mitigated with daily antigen testing

**DOI:** 10.1101/2021.03.03.21252838

**Authors:** Gage K. Moreno, Katarina M. Braun, Ian W. Pray, Hannah E. Segaloff, Ailam Lim, Keith Poulson, Jonathan Meiman, James Borcher, Ryan P. Westergaard, Michael K. Moll, Thomas C. Friedrich, David H. O’Connor

**Affiliations:** Department of Pathology and Laboratory Medicine, University of Wisconsin-Madison USA 53711; Department of Pathobiological Sciences, University of Wisconsin-Madison USA 53711; Wisconsin Department of Health Services, USA 53703; Epidemic Intelligence Service, Centers for Disease Control and Prevention USA 30333; Wisconsin Veterinary Diagnostic Laboratory, University of Wisconsin-Madison USA 53711; Department of Family Medicine, Division of Sports Medicine, Ohio State University USA 43210; Department of Medicine, University of Wisconsin-Madison, USA 53711; Athletic Department, University of Wisconsin-Madison USA 53711

**Author notes:** **Corresponding author contact information:** David O’Connor. These authors contributed equally (Gage K. Moreno, Katarina M. Braun).

**Keywords:** SARS-CoV-2, antigen testing

## Abstract

**Background:** High frequency, rapid turnaround SARS-CoV-2 testing continues to be proposed as a way of efficiently identifying and mitigating transmission in congregate settings. However, two SARS-CoV-2 outbreaks occurred among intercollegiate university athletic programs during the fall 2020 semester despite mandatory directly observed daily antigen testing.

**Methods:** During the fall 2020 semester, athletes and staff in both programs were tested daily using Quidel’s Sofia SARS Antigen Fluorescent Immunoassay (FIA), with positive antigen results requiring confirmatory testing with real-time reverse transcription polymerase chain reaction (RT-PCR). We used genomic sequencing to investigate transmission dynamics in these two outbreaks.

**Results:** In Outbreak 1, 32 confirmed cases occurred within a university athletics program after the index patient attended a meeting while infectious despite a negative antigen test on the day of the meeting. Among isolates sequenced from Outbreak 1, 24 (92%) of 26 were closely related, suggesting sustained transmission following an initial introduction event. In Outbreak 2, 12 confirmed cases occurred among athletes from two university programs that faced each other in an athletic competition despite receiving negative antigen test results on the day of the competition. Sequences from both teams were closely related and unique from strains circulating in the community, suggesting transmission during intercollegiate competition.

**Conclusions:** These findings suggest that antigen testing alone, even when mandated and directly observed, may not be sufficient as an intervention to prevent SARS-CoV-2 outbreaks in congregate settings, and highlights the importance of supplementing serial antigen testing with appropriate mitigation strategies to prevent SARS-CoV-2 outbreak in congregate settings.

**Summary:** High frequency, rapid turnaround SARS-CoV-2 testing continues to be proposed as a way of efficiently identifying and mitigating transmission in congregate settings. However, here we describe two SARS-CoV-2 outbreaks occurred among intercollegiate university athletic programs during the fall 2020 semester.

## Introduction

Timely reporting of SARS-CoV-2 test results is critical for controlling transmission through prompt public health action, yet at times during 2020, turnaround times for SARS-CoV-2 test results in the United States have averaged 4 days, with some individuals waiting 10 days or more [1]. While turnaround times in early 2021 have improved, the lag between specimen collection and receipt of a test result continues to represent a window in which the risk of viral spread from SARS-CoV-2-infected individuals is high. Rapid antigen tests, like Abbott’s BinaxNow COVID-19 Ag Card and Quidel’s Sofia SARS Antigen FIA, can reduce this lag between testing and results reporting [2–5]. Because of these qualities, high-frequency, rapid turnaround SARS-CoV-2 antigen testing has been proposed as a prevention strategy in many congregate settings where SARS-CoV-2 infection risk is elevated [6–8].

In data submitted for emergency use authorization, the Sofia SARS Antigen FIA antigen test reported a sensitivity of 97% and specificity of 100% when used for symptomatic patients within five days of symptom onset [9,10]. It therefore follows that serial antigen testing could rapidly identify persons with symptomatic infections enabling rapid isolation of such individuals [2,4]. Recent studies, however, have found that Sofia SARS Antigen FIA antigen was less sensitive (41.2% sensitivity) when individuals were asymptomatic [10–13]. Use of Sofia SARS Antigen FIA on asymptomatic patients is not included in the FDA authorization and is considered an “off-label” use of the test. Nonetheless, many universities and other congregate settings have used the tests for asymptomatic screening. The potential for false-negative antigen results among asymptomatic patients may present a significant risk in that a negative test could result in risk disinhibition behavior in a patient who may be infectious during their pre-symptomatic period, which could lead to sustained and increased viral spread[14] (**Figure 1**).

**Figure 1.**
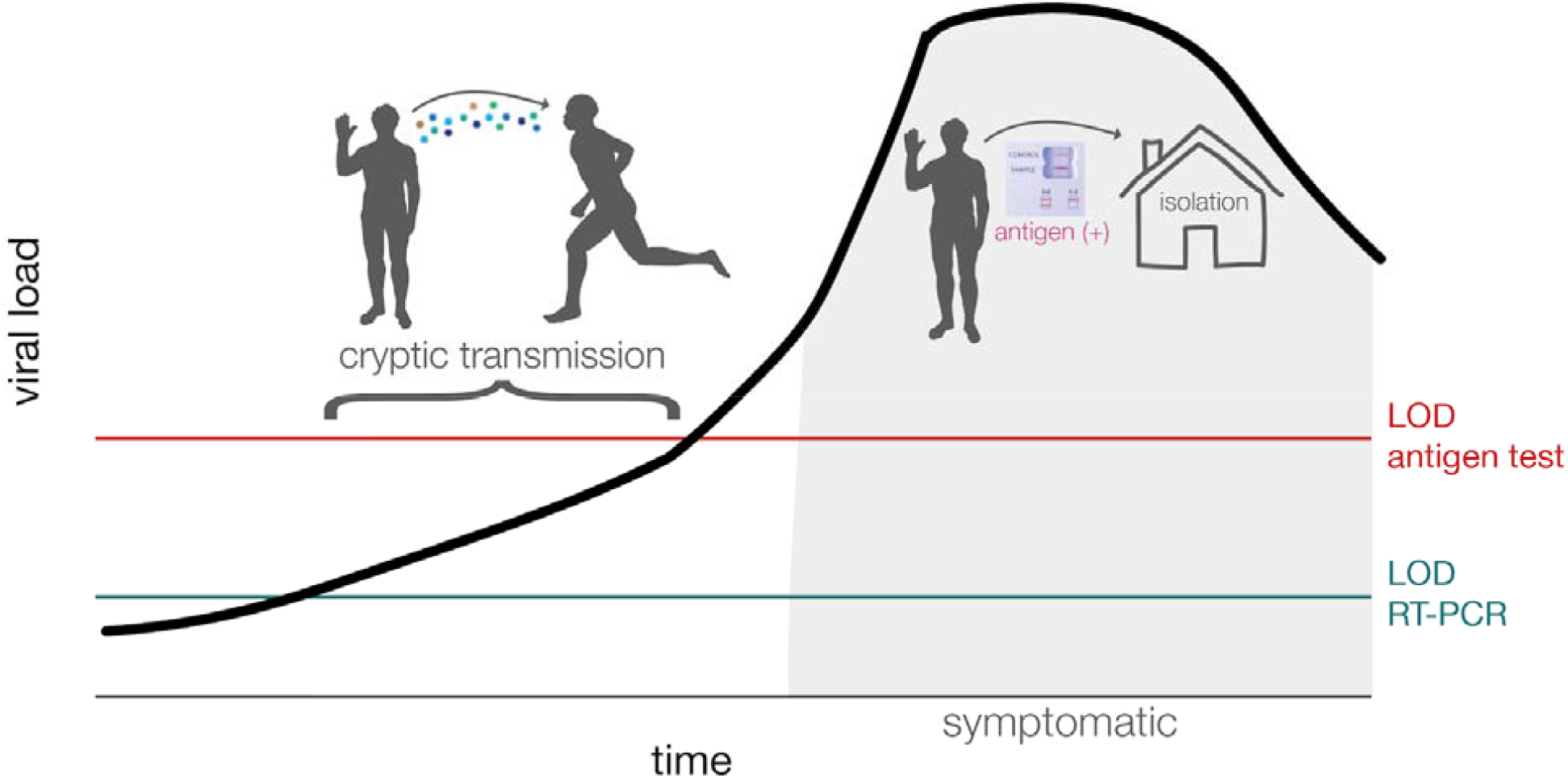
Graphical abstract of cryptic transmission that could occur when a person is asymptomatic and the amount of virus remains below the limit of detection for antigen tests despite the person being potentially infectious to others. This is a schematic and is meant to represent general, not quantitative, relationships among these variables. LOD = limit of detection.

## Methods

### A university implemented daily SARS-CoV-2 antigen testing for college athletics

The two outbreaks occurred among athletes and staff affiliated with a university’s intercollegiate athletics programs despite daily SARS-CoV-2 testing with Quidel’s Sofia SARS Antigen FIA. Both sports involved in the outbreaks were considered “high-risk” by the national collegiate athletics association (NCAA) due to frequent contact and collision between athletes during play. Students and staff affiliated with the two athletics programs began daily antigen testing for SARS-CoV-2 in September 2020. Daily antigen testing was not required for persons with a RT-PCR-confirmed SARS-CoV-2 infection in the past 3 months, and persons experiencing symptoms consistent with COVID-19, as symptomatic persons received RT-PCR testing without initial antigen testing. For remaining asymptomatic students and staff, antigen testing was conducted using anterior nasal swabs that were self-collected each morning under the direct supervision of a nurse. Antigen test results were provided to athletics department medical staff who coordinated exclusion from team activities and confirmatory testing, but were not reported back to students and staff.

A negative antigen result meant an individual could engage in all sport-related activities, like indoor meetings, practices, scrimmages, and intercollegiate competitions. Athletes and staff with positive antigen results were immediately excluded from team activities by department medical staff and subject to confirmatory testing with RT-PCR using the TaqPath COVID-19 Combo Kit (Thermo Fisher Scientific). Students and staff with positive RT-PCR results were excluded from team activities for 21 days and were interviewed by university staff to identify close contacts. Close contacts of RT-PCR-confirmed students or staff were required to self-quarantine for 14 days from the date of last contact per public health guidance [17]. Importantly, contact tracing for student athletes did not include contacts that occurred during practices, competitions, meetings, or other team activities, but could include contacts that occurred during social activities or at home (e.g., roommates). In addition to daily antigen testing, the athletic programs implemented a physical distancing policy requiring all students and staff to be at least six feet apart during meetings, and mandatory mask use during team activities.

### Epidemiological investigation

Confirmed cases of COVID-19 were defined as students or staff affiliated with the two athletics programs who received a positive SARS-CoV-2 RT-PCR result during the outbreak period. False negative antigen results were defined as a negative antigen test result and a positive RT-PCR result that were collected on the same day. During each outbreak, once the number of confirmed cases reached the threshold established by intercollegiate athletics conference protocols, in-person team activities were suspended, and all students and staff were tested with RT-PCR. Specimens that tested positive by RT-PCR confirmation were used for sequencing analysis.

The names of universities, the specific sports, and relevant dates have been removed from the report to protect the privacy of the students and staff involved. We used identifiers (Athletics-##) to denote individuals associated with these outbreaks. Dates are encoded as X-day-YY, ‘X’ indicates the outbreak investigated, and ‘YY’ indicates the day of that outbreak. The first notable event for each outbreak is “day 0” – in Outbreak 1, this was a negative antigen test for the index case (who later tested positive by RT-PCR), and in Outbreak 2, this was the date of the first competition between the two teams. This activity was reviewed by CDC and was conducted in a manner consistent with applicable federal law and CDC policy^1^.

### Laboratory Methods

We obtained a waiver of HIPAA Authorization and were approved to obtain the clinical samples along with a Limited Data Set by the Western Institutional Review Board (WIRB #1-1290953-1). Sequences for this study were derived from 36 total nasopharyngeal (NP) swab samples collected from Outbreak 1 (n=32) and Outbreak 2 hosting team (n=5), as well as the visiting team’s samples in Outbreak 2 (n=5).

### Outbreak 1 viral RNA isolation

Nasal swabs were collected and placed in 3mL phosphate buffered saline. RNA was extracted from 190 uL of sample using the MagMAX™ Viral/Pathogen II (MVP II) Nucleic Acid Isolation Kit (Thermo Fisher Scientific, Waltham, MA) and eluted in a volume of 50 µL according to manufacturer’s instructions. 5µL of RNA was quantitated using a one-step RT-PCR using a TaqPath COVID-19 Combo Kit (Thermo Fisher Scientific).

### Outbreak 2 viral RNA isolation

Nasopharyngeal swabs were received in 3mL of transport medium (VTM). Viral RNA (vRNA) was extracted from 100□μl of VTM using the Viral Total Nucleic Acid Purification kit (Promega, Madison, WI, USA) on a Maxwell RSC 48 instrument, using manufacturer guidelines, and was eluted in 50 μL of nuclease free H_2_O.

### Complementary DNA (cDNA) generation

Complementary DNA (cDNA) was synthesized using a modified ARTIC Network approach[15]. Briefly, vRNA was reverse transcribed with SuperScript IV Reverse Transcriptase (Invitrogen, Carlsbad, CA, USA) using random hexamers and dNTPs, according to manufacturer’s guidelines.

### Multiplex PCR to generate SARS-CoV-2 genomes

SARS-CoV-2-specific multiplex PCR for nanopore sequencing was performed, similar to amplicon-based approaches as previously described[15,16]. In short, primers for 96 overlapping amplicons spanning the entire genome with amplicon lengths of 500 bp and overlapping by 75 to 100 bp between the different amplicons were used to generate cDNA. cDNA (2.5□μL) was amplified in two multiplexed PCR reactions. Following amplification, samples were pooled together before ONT library preparation.

### Library preparation and sequencing

A total of 5ng for each sample was made compatible for deep sequencing using the one-pot native ligation protocol with Oxford Nanopore kit SQK-LSK109 and its Native Barcodes (EXP-NBD104 and EXP-NBD114)[15]. Up to 24 samples were pooled prior to being run on the appropriate flow cell (FLO-MIN106) using the 72hr run script.

### Processing raw ONT data

Sequencing data was processed using the ARTIC bioinformatics pipeline (https://github.com/artic-network/artic-ncov2019)[17]. Consensus sequences were assembled for samples with greater than 400x coverage. Samples were excluded from analysis if gaps in the consensus sequence totaled ≥20% of the genome. The entire ONT analysis pipeline is available at https://github.com/gagekmoreno/SARS-CoV-2-in-Southern-Wisconsin.

### Phylogenetic analysis

Phylogenetic analysis was completed using tools implemented in Nextstrain custom builds (https://github.com/nextstrain/ncov)[18,19]. Time-resolved and divergence phylogenetic trees were built using the standard Nextstrain tools and scripts[18,19]. We used custom python scripts to filter and clean metadata. Sequences names were coded as OB#-T#-A#. Where OB signifies the outbreak, T represents the team that the sequence came from, and A is the athlete from which the sample that the sequence was derived originated.

### Data availability

Source data after mapping to SARS-CoV-2 reference genome (Genbank: MN908947.3) have been deposited in the Sequence Read Archive (SRA) under bioproject PRJNA614504.

## Results

### Outbreak 1: Cases can be linked to a single viral introduction

An athlete (Athletics-1) received a negative antigen test result the morning of day-0. Later that day, Athletics-1 attended an indoor meeting with approximately 10 other student-athletes and staff in which attendees reportedly sat six feet apart and wore masks at all times. The following morning (day-1), Athletics-1 received a positive antigen test result followed by RT-PCR swab for confirmation. The confirmatory RT-PCR result was positive with a Ct of 15.9 and the athlete began experiencing symptoms by mid-afternoon of day-1. During day-3 through day-7, four attendees of the initial day-0 meeting developed symptoms and received subsequent positive RT-PCR results. Additionally, three roommates of Athletics-1 who did not attend the meeting developed symptoms on day-4 and received positive RT-PCR results on day-4 and day-5. In-person team activities were suspended on day-8 to prevent additional transmission.

Program-wide RT-PCR and antigen testing was conducted seven times throughout the outbreak period (day-7, 10, 13-17, and 20). Mass RT-PCR testing identified 21 new SARS-CoV-2 infections among students and staff. Of these, 18 (86%) were negative on contemporaneous rapid antigen tests. Among 11 positive antigen results obtained during mass testing, 4 (36%) were confirmed with RT-PCR and 7 (64%) received negative RT-PCR results.

Overall, during Outbreak 1, 32 individuals (22 students and 10 staff) from the program had laboratory-confirmed SARS-CoV-2 infections (**Figure 2a**). Of confirmed cases, 4 (13%) were tested with RT-PCR because they were symptomatic, 7 (22%) were antigen-positive and received RT-PCR confirmation, and 21 (66%) were positive during mass RT-PCR testing. Contact tracing interviews found that 13 (40%) of 32 confirmed cases attended a team meeting where someone with confirmed COVID-19 was present and in their infectious period; 6 (13%) had close contact with a roommate with COVID-19; and 8 (25%) did not have any documented exposures (**Table 1)**.

**Table 1.**
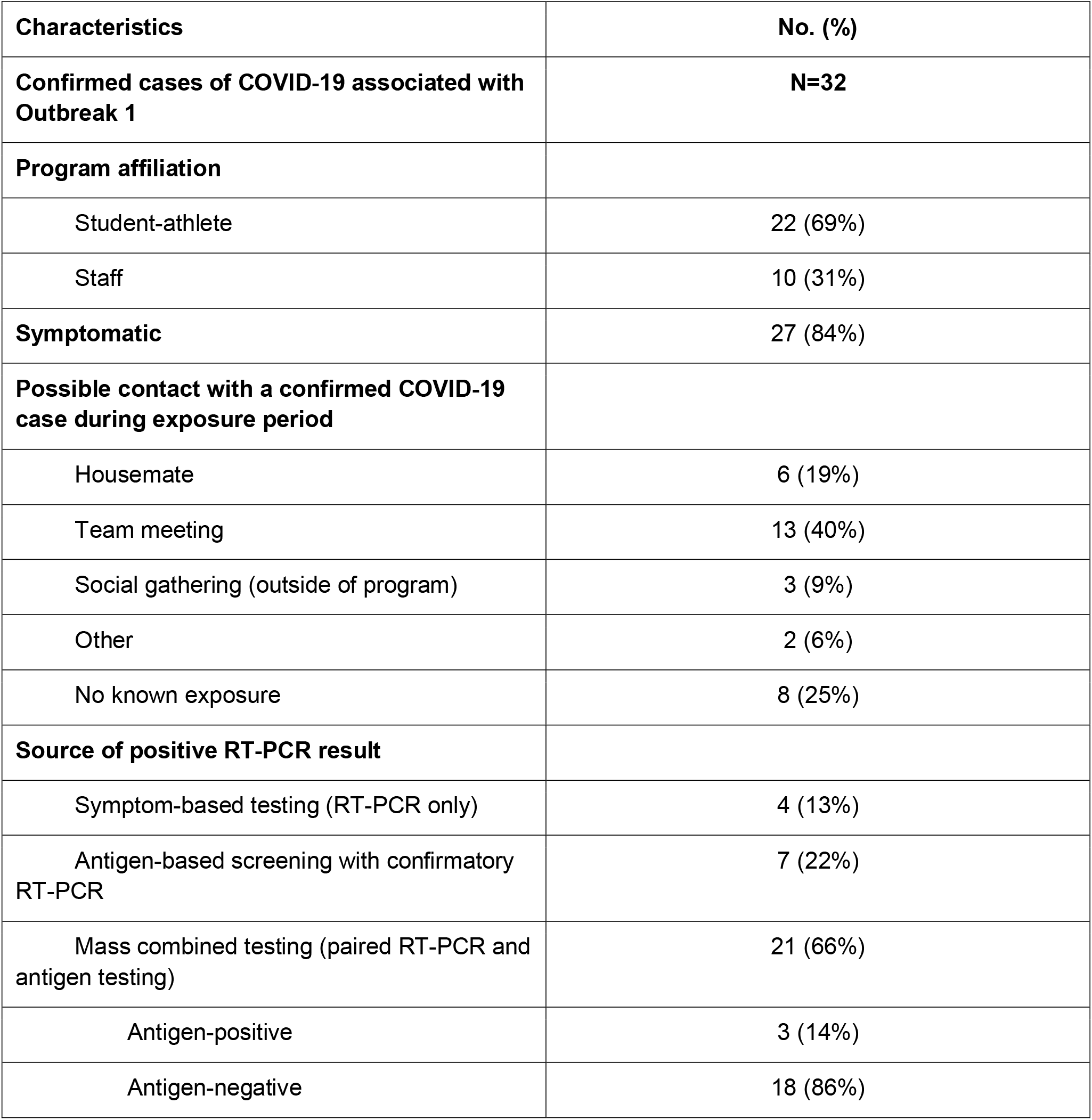
Characteristics and exposure details for confirmed COVID-19 cases (n=32) during Outbreak 1.

| Characteristics | No. (%) |
| --- | --- |
| **Confirmed cases of COVID-19 associated with Outbreak 1** | **N=32** |
| **Program affiliation** |  |
| Student-athlete | 22 (69%) |
| Staff | 10 (31%) |
| **Symptomatic** | 27 (84%) |
| **Possible contact with a confirmed COVID-19 case during exposure period** |  |
| Housemate | 6 (19%) |
| Team meeting | 13 (40%) |
| Social gathering (outside of program) | 3 (9%) |
| Other | 2 (6%) |
| No known exposure | 8 (25%) |
| **Source of positive RT-PCR result** |  |
| Symptom-based testing (RT-PCR only) | 4 (13%) |
| Antigen-based screening with confirmatory RT-PCR | 7 (22%) |
| Mass combined testing (paired RT-PCR and antigen testing) | 21 (66%) |
| Antigen-positive | 3 (14%) |
| Antigen-negative | 18 (86%) |

**Figure 2.**
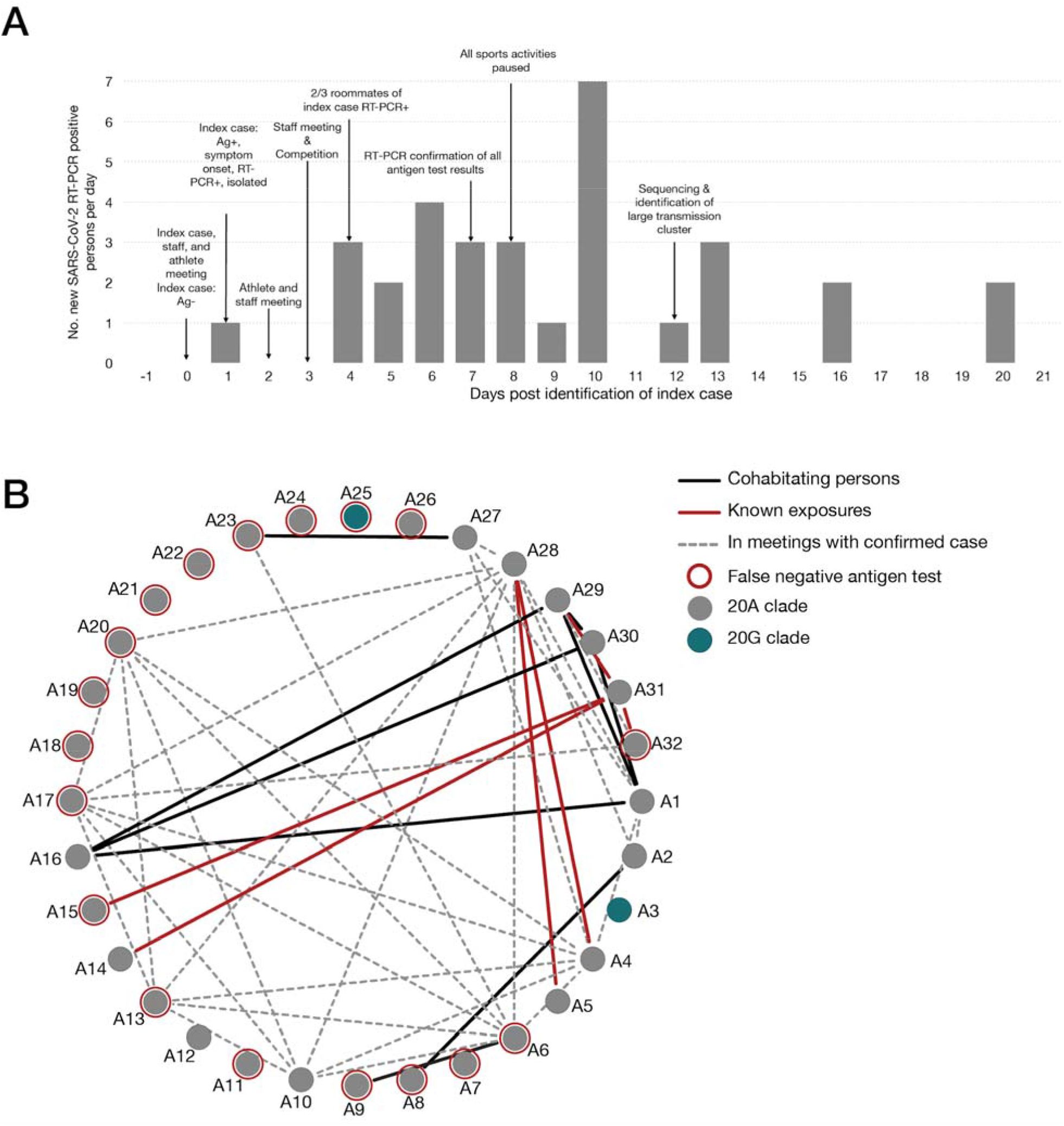
Overview of Outbreak 1. A) Epidemic curve for confirmed COVID-19 cases (n = 32) among students and staff associated with the athletics program during Outbreak 1. Abbreviations: Ag = antigen; COVID-19 = coronavirus disease 2019; RT-PCR = reverse transcription–polymerase chain reaction. B) Graphical representation of known interactions between all persons in the athletics program affected by Outbreak 1. Roommates are shown by a solid black line; confirmed close contact with a positive case as identified through contact tracing interviews is shown with a red line; r persons who attended indoor team meetings together while following physical distancing policies (> 6 feet apart and wearing masks) are shown with a dashed gray line; persons who received false negative antigen results are shown with a red circle.

To investigate the relationship among SARS-CoV-2 cases in outbreak 1, we generated consensus sequences for 26 (81%) of 32 RT-PCR positive samples using the ARTIC Network tiled amplicon approach on an Oxford Nanopore Technologies MinION [15,16]. Samples from the remaining six RT-PCR test-positive individuals in Outbreak 1 were not available at the time of sequencing and were excluded from this analysis. We found that 24 (92%) of these 26 genomic sequences cluster tightly in the Nextstrain 20A clade on a time-resolved tree and are separated by 0-2 fixed consensus nucleotide differences (**Figure 3**). The limited diversity of viruses detected in the 24 individuals suggests sustained transmission of SARS-CoV-2 following a single introduction [20–22].Viruses from Athletics-3 and Athletics-26 did not appear to be part of the primary transmission cluster. As of 1-day-40, there was no evidence for onward spread within the program originating from Athletics-3 or Athletics-26. The viruses infecting these individuals cluster more closely with sequences seen in the community.

**Figure 3.**
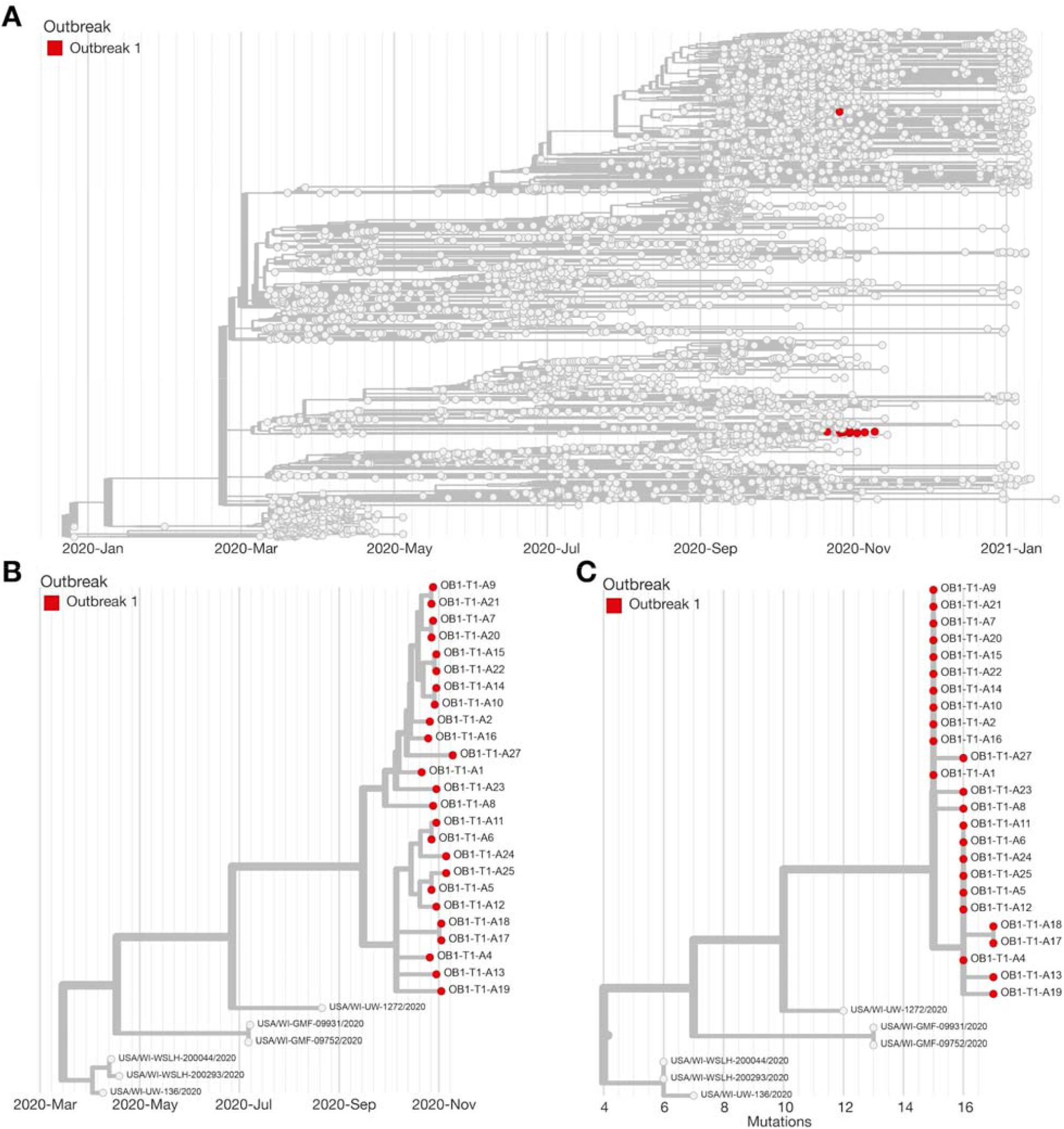
Phylogeny of Outbreak 1. A) Time-resolved phylogenetic tree created using Nextstrain tools and nomenclature showing the team sequences contextualized with all available community sequences (gray) for 25 (78%) of 32 confirmed cases associated with Outbreak 1; tips affiliated with Outbreak 1 are colored red. B) Zoomed in time-resolved phylogeny showing all of these samples are part of the same athletics cluster. C) Divergence tree showing the number of mutations each sequence has relative to Wuhan/WH01/2019 (Genbank: MN908947.3), a standard reference comparison sequence.

### Outbreak 2: SARS-CoV-2 transmission during intercollegiate competition

Two teams from different universities engaged in intercollegiate competitions on consecutive days (day-0 and day-1). Both teams underwent daily antigen testing and received all negative antigen results in the week preceding the competitions, including both competition days (day-0 and day-1). No testing was conducted on day-2. On day-3, an athlete from Team 2 received a positive antigen test result, which was confirmed by RT-PCR. No athletes or staff on either team were quarantined from contact with the index athlete that occurred during competition on day-0 and day-1. During day-5 through day-10, multiple athletes on both teams developed symptoms and received positive antigen and RT-PCR results. On day-6, all athletes on Team 1 were tested by RT-PCR and in-person team activities were suspended. Overall, 12 athletes (seven from Team 1 and five from Team 2) had confirmed SARS-CoV-2 infections during this outbreak (**Figure 4**).

**Figure 4.**
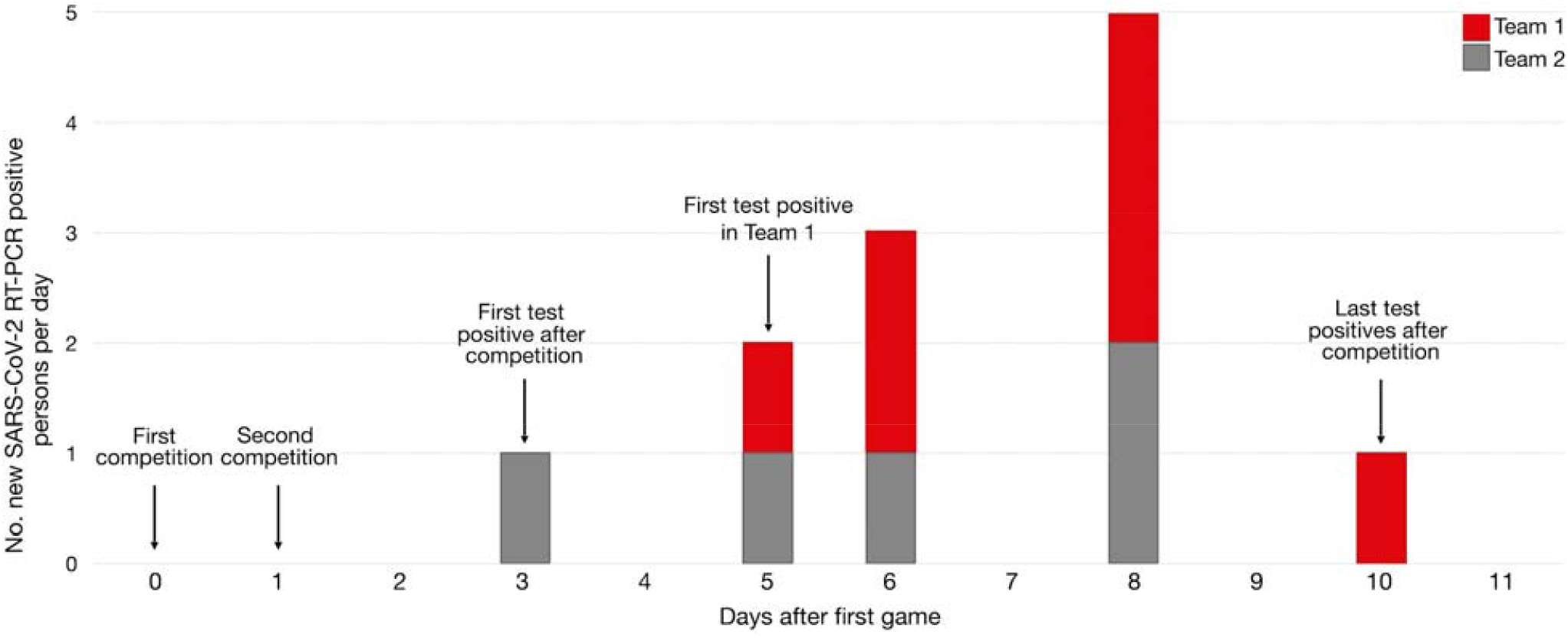
Overview of Outbreak 2. A) Epidemic curve for Outbreak 2 showing confirmed (n = 12) COVID-19 cases within the two intercollegiate teams. Testing was not conducted on day-2.

**Figure 5.**
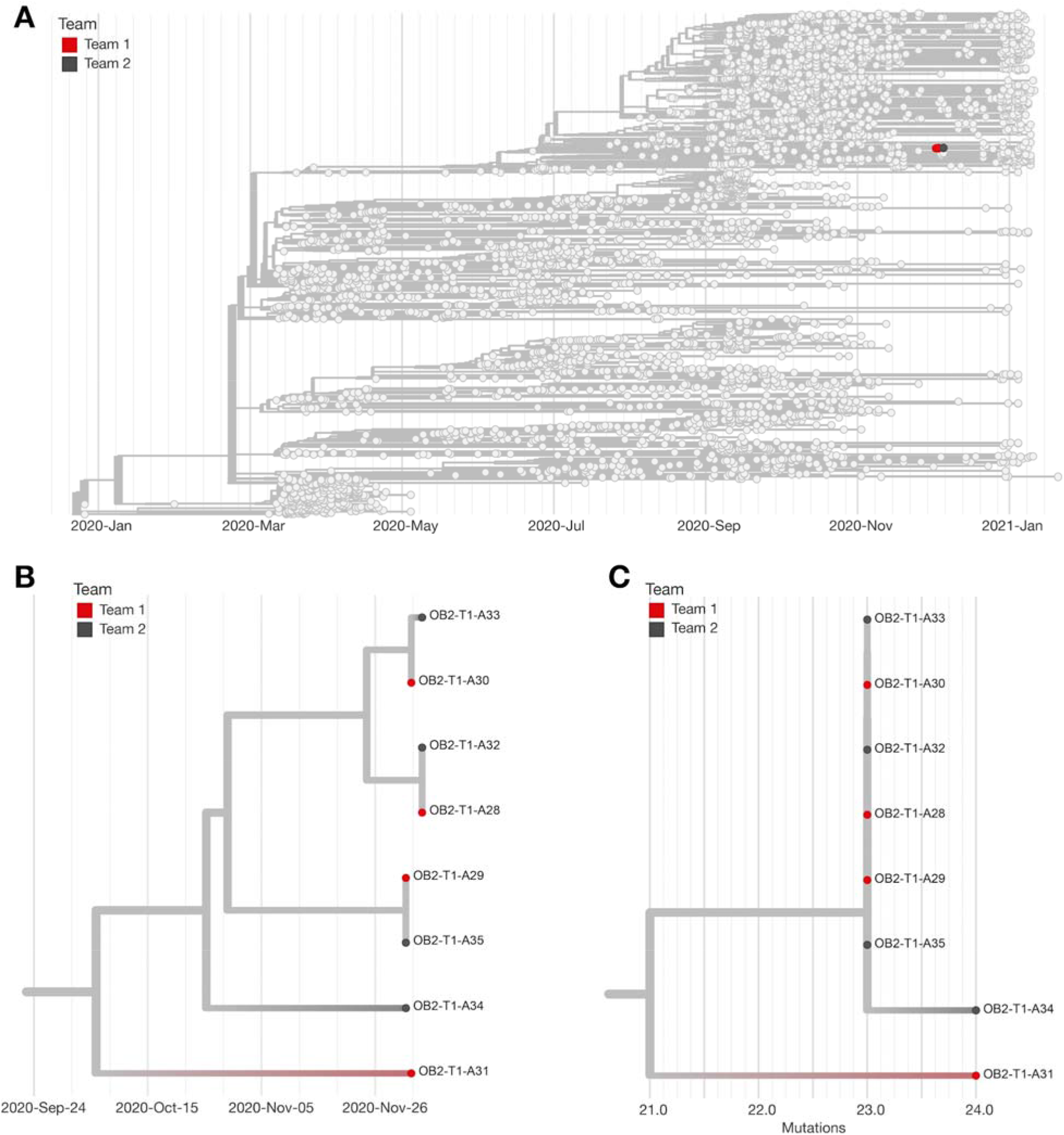
Phylogeny of Outbreak 2. A) Time-resolved phylogenetic tree created using Nextstrain tools and nomenclature showing 8 (67%) of 12 available samples from Outbreak 2 sequences contextualized with all available community sequences (light gray). Tips affiliated with Team 1 are colored red, and Team 2’s sequences are colored dark gray. B) Zoomed in time-resolved phylogeny showing all these samples are part of the same athletics cluster. C) Divergence tree showing the number of mutations each sequence has relative to Wuhan/WH01/2019 (Genbank: MN908947.3), a standard reference comparison sequence.

To determine whether the source of these infections could be linked to competition despite negative antigen results on the day of competition, we generated eight consensus sequences from ten available samples. All eight virus sequences (four from each team) clustered tightly in the 20G clade on a time-resolved tree and were separated by 0-2 fixed consensus nucleotide differences (**Figure 4**). Given the known epidemiological associations between these teams, this likely represented a single transmission cluster [20–22].

The genetic sequences of the viruses infecting the individuals in Outbreak 2 were distinct from the viruses circulating within the community where Outbreak 2 occurred. Moreover, sampling of individuals in Outbreak 2 revealed a unique mutation, encoding Spike P26Y, that was present in viral sequences from the samples from both teams but was not otherwise seen in the larger community where Team 1 was located. Given the depth of surveillance community sequencing in Team 1’s community available during the outbreak period (∼4.7% of test-positive cases), it is unlikely that this unique signature arose independently in the community where Team 1 is located.

## Discussion

The SARS-CoV-2 testing strategy of daily, directly observed, rapid antigen testing implemented by intercollegiate athletics programs nationwide has been resource-intensive, yet its impact on SARS-CoV-2 transmission in this setting has not been evaluated. In this report, we described two outbreaks within intercollegiate athletics programs in which daily antigen testing was unable to interrupt SARS-CoV-2 transmission. In Outbreak 1, the index athlete received a positive RT-PCR results with a low Ct value less than 24 hours after testing negative by the Sofia SARS Antigen FIA. This individual was likely infectious on the date of the negative antigen test, as four other individuals in the day-0 meeting contracted SARS-CoV-2 over the ensuing week. Sustained transmission within the program followed when additional exposures from presymptomatic and undetected SARS-CoV-2 infections occurred – at least 13 of the 32 outbreak-associated cases attended team meetings with individuals who had received negative antigen results yet were in their infectious period.

Transmission within the program was not interrupted until the program implemented serial RT-PCR testing, a strategy that led to identification of 21 new confirmed SARS-CoV-2 infections, 18 of which were negative on contemporaneous antigen tests. Our findings suggest that serial antigen testing as a control strategy may have limited sensitivity for detecting early asymptomatic infections, and that prevention of future outbreaks in these settings may require a combination of more sensitive molecular tests (e.g., RT-PCR) and improved mitigation measures.

Contact tracing during Outbreak 1 identified interactions between individuals that may have contributed to at least 21 (66%) of the 32 confirmed cases (**Figure 2b**). These interactions represent multiple breaches of the university’s mitigation strategy and combined with the limitations of the antigen testing protocol, resulted in sustained person-to-person viral spread throughout the team. In particular, the team continued to have physically distanced (6 feet apart) in-person meetings with cloth masks until all in-person team activities were suspended to prevent further spread. Per public health and university guidelines, attendees in these meetings were not quarantined, a step that may have prevented onward transmission during this outbreak. Roommates and household contacts of student-athletes could represent additional sources of infection in Outbreak 1. In some cases, housemates of infected team members were not required to quarantine due to the large size of the house and the university’s assessment that physical distancing was achievable in this area. Continuing indoor in-person meetings and not quarantining potential contacts represent possible breaches in university’s SARS-CoV-2 mitigation plan that, combined with the limitation of antigen testing, permitted viral spread throughout the team in Outbreak 1.

In Outbreak 2, we used genomic sequencing to demonstrate that SARS-CoV-2 transmission likely occurred between two teams during athletic competition despite both teams receiving negative antigen results prior to competition. Supporting evidence for inter-collegiate transmission included detection of a unique mutation, Spike P26Y, that was common to the samples from both teams but not otherwise seen in the community where Team 1 is located. Given these findings, the most parsimonious explanation is that an infection acquired in the community by the index athlete on Team 2 was transmitted to other individuals on both teams during the time of competition.

The potential for inter-collegiate transmission during an athletic competition has important implications for SARS-CoV-2 serial testing strategies and in-competition mitigation protocols. First, antigen testing on the competition dates failed to identify the index case, who may have been infectious and exposed other athletes. Like Outbreak 1, more sensitive molecular tests may have identified the source case and allowed for exclusion from the competition. Second, this investigation shows that athletic competition may pose a risk for SARS-CoV-2 transmission, particularly in sports where direct physical contact occurs. This outbreak occurred during an athletic competition that included contact and collision, and is considered “high-risk” by NCAA. Despite the short duration of contact between athletes, transmission risk can be exacerbated by heavy breathing and shouting without masking, which regularly occurs in this sport and has been associated with SARS-CoV-2 outbreaks in other athletics competitions [30].

The findings in this report are subject to several limitations. First, we were not able to perform genomic sequencing on all positive samples from these outbreaks (34 of 44 samples were sequenced) either because of high Ct values on RT-PCR or lack of sample availability. Second, contemporaneous antigen and RT-PCR samples in Outbreak 1 were not collected as “paired” swabs (simultaneous swabbing of two nares) and may not be comparable to other antigen test evaluations. Similarly, the performance of antigen tests in this context of daily serial testing measured their ability to identify early presymptomatic infections, and may not be generalizable to antigen test performance in other settings. Third, our ability to determine the source of infections in these outbreaks was limited by incomplete contact tracing data. Undocumented exposures between athletes and staff may have occurred outside of organized team activities that could have caused infections that were attributed to team meetings or in-competition transmission; although the strength of genomic clustering and epidemiologic evidence from these investigations suggests that such occurrences were rare.

Among athletics programs and other congregate settings where outbreaks may spread rapidly after introduction of SARS-CoV-2, serial antigen testing alone may not be sufficient to prevent outbreaks. A robust testing strategy should be supplemented with multilayered prevention strategies that includes correct and consistent mask use, physical distancing, increased hand hygiene and disinfection, avoiding crowds and poorly ventilated spaces, and isolation of symptomatic individuals regardless of antigen test result[13,23–25]. Serial testing with RT-PCR may identify additional cases that were not detected by antigen testing, but the increased sensitivity would have to be balanced with laboratory resources and increased turnaround times.

## Data Availability

https://www.ncbi.nlm.nih.gov/bioproject/?term=PRJNA614504

## Acknowledgments

We gratefully acknowledge Dr. Trevor Bedford and the entire Nextstrain team for making Nextstrain phylogenetic tools publicly available and for their commitment to tracking the global spread of SARS-CoV-2. We also acknowledge the GISAID team for maintaining the largest public repository of SARS-CoV-2 sequence- and metadata.

## Footnotes

1 See e.g., 45 C.F.R. part 46.102(l)(2), 21 C.F.R. part 56; 42 U.S.C. §241(d); 5 U.S.C. §552a; 44 U.S.C. §3501 et seq.

